# Multi-center improvement in screening for pain that affects activities in adults with cerebral palsy

**DOI:** 10.1101/2025.01.05.25319957

**Authors:** Amy F. Bailes, Garey H. Noritz, Duncan O. Wyeth, Elizabeth J. Lucas, Elisabeth B. Bates, Hana Azizi, Cristina A. Sarmiento, Deborah E. Thorpe, Stephen A. Nichols, Jodi Kreschmer, Stephen Wisniewski, Mary Gannotti, Cerebral Palsy Research Network

**Affiliations:** Cincinnati Children’s Hospital Medical Center Division of Occupational therapy and Physical Therapy, Division of Patient Services Research, University of Cincinnati College of Medicine, Cincinnati, OH 45229; Nationwide Children’s Hospital, Complex Health Care Program; 700 Children’s Drive, Columbus, OH 43205; Mohawk, Michigan 49950. Member Cerebral Palsy Research Network-- Community Action Committee; Columbia University Medical Center, New York, NY 10032; University of Colorado Anschutz, Department of Physical Medicine and Rehabilitation; 13123 East 16th Avenue, Box 285, Aurora CO, 80045; University of North Carolina, Chapel Hill, Department of Health Sciences, Division of Physical Therapy; Bondurant Hall, CB #7135, Chapel Hill, NC 27599; Mt. Washington Pediatric Hospital, 1708 W. Rogers Avenue, Baltimore, MD 21209; University of Michigan Medical School, Department of Physical Medicine and Rehabilitation; 325 E Eisenhower Parkway Suite 100, Ann Arbor, MI 48108; University of Pittsburgh, Epidemiology Data Center, 4420 Bayard Street, Pittsburgh, PA 15260; Department Rehabilitation Sciences, University of Hartford, 200 Bloomfield Avenue, West Hartford, CT 06117

**Keywords:** Adults, cerebral palsy, pain, quality improvement, learning health network

## Abstract

**Purpose:** Increase screening for pain in adults with cerebral palsy (CP) across three centers and examine factors associated with pain that affect activities.

**Materials and Methods:** Using the quality improvement (QI) infrastructure of the Cerebral Palsy Research Network (CPRN), we implemented interventions to improve screening at clinic visits for pain that affects activities for adults with CP. Three physicians from two CPRN centers performed interventions August 2021- June 2023 before spreading to a fourth physician at a third CPRN center October 2022. To track progress, we collected visit data cross sectionally every two weeks. Descriptive statistics, analysis of variance, and logistic regression evaluated relationships in a sample cohort of all visits after screening practices had been established.

**Results:** Screening improved from 42% at baseline to over 90%. After three months of sustained screening, we assessed 423 unique visits. Pain was reported at 185/423 (44%) of the visits. Of the 185 with pain reported, 100 (54%) reported pain that affected activities. Increasing age, female gender, and motor function were associated with pain (p<.001) and pain that affects activities (p<.01). Females reported pain 3.4 and pain that affects activity 2.2 times more than males.

**Conclusion:** QI methodology was successful at improving screening for pain that affects activities in adults with CP at clinic visits. Lower rates of pain were found (44%) than previous reports, with similar findings about pain affecting activities and associated characteristics. We propose continued screening with improvement in differentiating proxy vs self-report and including other domains of pain important to guide care such as location and chronicity.

## INTRODUCTION

The number of adults with cerebral palsy (CP) is increasing with global demographic trends of an aging population.[1] Although CP is the most common physical disability in children,[2] most of the population is now over the age of 18. [3] CP impacts everyone differently in terms of type and magnitude of motor impairment and other co-occurring conditions.[4] However, as a group, adults with CP have an increased risk of poor health outcomes compared to peers.[5–8] Initiatives to improve outcomes for adults with CP should be a priority for providers, policy makers, and clinicians given the intensity of resources needed to manage secondary conditions resulting from lack of mobility and other predisposing risk factors.[1, 9]

One priority identified by adults with CP, clinicians, and researchers is improved identification and management of pain.[10] Current estimates for pain prevalence among adults with CP are about 70%.[11] An international systematic review identified women at significantly higher risk for pain than men and those classified as Gross Motor Function Classification System (GMFCS)[12] II and IV are at higher risk compared to individuals classified as GMFCS I. Screening for pain among adults with CP is recommended given the high prevalence and costly consequences.[11] There was a limited amount of data from the US in the review,[11] as currently there exists no standardized way providers document pain and patient characteristics, nor is there a mechanism to aggregate and analyze data.

Although pain screening is part of care guidelines in the US, *standardization* of screening for pain during clinic visits with adults with CP among providers is a first step for developing effective management and treatment protocols.[13] The 2018 updated Joint Commission R3 Report Issue 11: *Pain Assessment and Management Standards for Hospitals* states that accredited institutions should improve pain assessment by concentrating on how pain is affecting patients’ physical function.[13] To date, there are no published descriptions of how medical centers are implementing standardized screening for pain and how it affects function among adults with CP.

Quality improvement (QI) methods would offer a systematic way to develop, test and implement changes to improve outcomes[14] for adults with CP. QI utilizes multiple “Plan, Do, Study, Act” (PDSA) cycles to standardize processes and structure to reduce variation, achieve predictable results, and improve outcomes for patients, healthcare systems, and organizations.[14]

The Cerebral Palsy Research Network (CPRN), established in 2015, includes multiple institutions engaging stakeholders to improve the health of individuals with CP. Like other Learning Health Networks,[15, 16] a core goal of the network is to engage in QI initiatives with the potential for impacting large numbers of consumers. The CPRN Adult Quality Improvement Community of Practice (QI CoP) includes clinicians, researchers, and community members. Screening for pain that affects activities was identified as a top priority by the QI CoP.

QI methods and CPRN QI CoP may provide a sustainable path towards improving practice and advancing care for pain in adults with CP. The purpose of this report was to improve screening for pain in adults with CP across three centers and examine factors associated with pain that affects activities in a cohort.

## METHODS

### Setting and Context

The QI project was initiated in August 2021 and reported through June 2023 at select CP outpatient clinics across three participating CPRN academic medical centers that serve adults (Nationwide Children’s Hospital (NCH), Columbia University Irving Medical Center (CUIMC), and University of Colorado Hospital (UCH)). The QI CoP team consisted of two developmental pediatricians, one nurse practitioner, two physical medicine and rehabilitation physicians, an expert in QI methodology, three rehabilitation experts/consultants, and two community members with CP. Three physicians in the QI CoP (two developmental pediatricians (NCH) and one physiatrist (CUIMC)) provided baseline data and participated in implementing interventions in their clinic setting. The QI CoP met twice a month to develop key drivers, design interventions, and evaluate a series of “Plan, Do, Study, Act” cycles.[14] The QI CoP members planned changes, observed results, and acted on what was learned. Each center’s Institutional Review Board (IRB) reviewed the QI project, and no IRB oversight or consent was required. The QI CoP used Microsoft Teams to manage de-identified electronic medical record (EMR) data extractions regularly. The Revised Standards for Quality Improvement Reporting Excellence (SQUIRE 2.0)[17] and Strengthening the Reporting of Observational studies in Epidemiology (STROBE)[18] were used to guide the information provided in this report.

### Intervention Methodology

The Model for Improvement,[14] was used to guide the project, At the start, the QI CoP team identified a global aim to improve quality of life for adults with CP. The QI CoP team envisioned a standardized process for assessing pain that affects activities and developed a Specific, Measurable, Actionable, Realistic, and Timely (SMART) aim to increase the percentage of adult visits (18 years or older) screened for pain that affects activities from 42% to 80%. QI CoP members with lived experience (DW, JK) shared that they always live with pain and are not concerned until it interferes with function. For this reason, they felt we should prioritize any pain and not differentiate between acute or chronic pain in this project. This is similar to Van Der Slot’s [11] systematic review where pain was dichotomized into no pain or any pain without subclassifying into acute or chronic. Further we did not differentiate between self-report or proxy. We intentionally focused on what we thought would be realistic during usual clinic visits based on input from the clinicians on the QI CoP. This aligns with QI methodology of testing interventions incrementally in order to make sustainable practice changes.[14]

Upon review of the literature, the QI CoP operationally defined screening for pain that affects activities with the pain attribute of the Health Utility Index 3 (HUI3).[19] The HUI3 pain attribute has five levels of responses that describe the severity of pain as it relates to limitations to normal daily activities including; 1) no pain, 2) pain that does not affect activities, 3) pain that prevents a few activities, 4) pain that prevents some activities, and 5) pain that prevents most activities. The HUI3 was originally designed to evaluate the health-related quality of life for very low birth weight infants, has been used with groups of patients having a wide range of conditions, and is valid for use by proxy among groups with impaired cognition.[20] Although it has not been validated specifically for use in CP,[21] it has been used successfully to identify pain, motor function, and health-related quality of life among children with CP, and to identify that increasing age and pain negatively predict quality of life in young adults with CP.[22]

The team developed a key driver diagram (KDD), which is a visual representation of the theory for improvement,[14] to guide and test interventions to drive improvement. The KDD was revised as the project progressed. The final KDD is shown in Figure 1. Change concepts – an approach to change that prompts new ways of thinking about how to improve the process – were used to brainstorm and organize the interventions. Change concepts[14] used in this project include “Change the work environment,” “Manage variation,” and “Give people access to information.”

**Figure 1:**
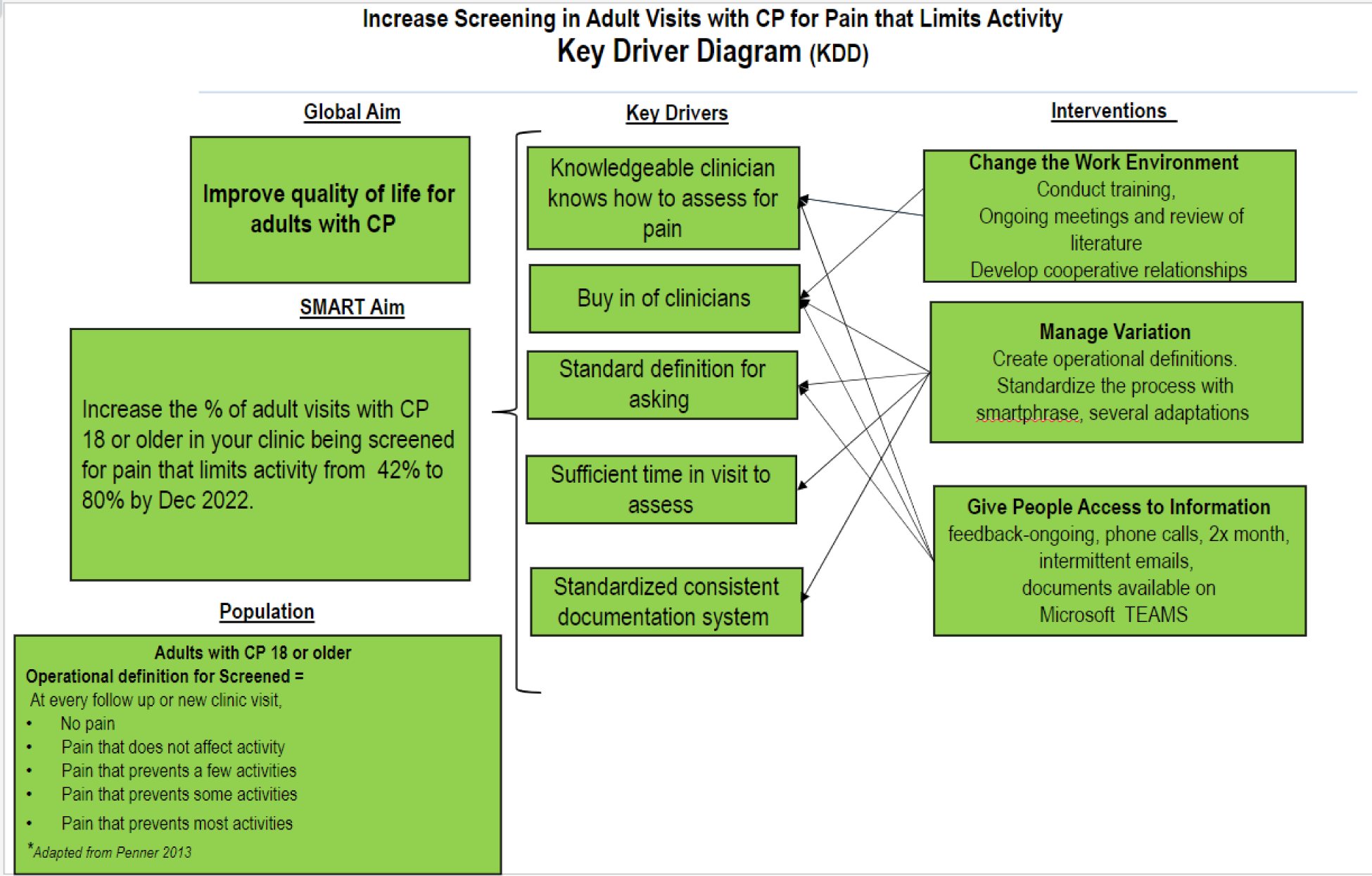
Final Key Driver Diagram. A diagram with three panels from left to right displaying in words the global and smart aims of the project and population on the far left, the key drivers in the center and the interventions used in the study to drive improvement in the far-right column. Arrows from the right column pointing to the middle column show which intervention affects which driver. A bracket is used from the middle column to the left side showing that the key drivers point towards improvement in the aim.

### Baseline Phase Methods

A retrospective chart review of all visits of adults with CP with the three participating QI CoP providers (2 developmental pediatricians at NCH, one PMR at CUIMC) was completed to establish the baseline rate (5/17/21-8/20/21) of screening for pain that affects activities. Although each center may have been asking about pain that affects activities during usual care, there was no standardized way to document this, and there was no way to measure if providers agreed on how to ask/classify responses.

### Testing Interventions

Testing the interventions started on 8/23/2021. The three providers or designated member of their clinical team were responsible for entering data at regular intervals into the secure database as close to real time as possible. Data was checked for completeness and accuracy by the study team monthly during regular meetings. Interventions tested during this phase and represented in the KDD were categorized according to the three following change concepts:

#### Change the Work Environment

The QI CoP participated in four training webinars offered by CPRN December 2020-March 2021. Also, reviews of QI topics occurred at regularly scheduled meetings. PDSA testing to change the work environment began with a literature review and discussions about pain, screening for pain in adults with CP, and raising awareness for the need to screen for pain that affects activities. During regularly scheduled meetings, we developed collaborative relationships with each other and strengthened members’ motivation for change.

#### Manage Variation

To manage variation the team agreed to standardize how “Screened for pain that affects activity” would be accomplished using terms included in the pain attribute of the HUI3. A smart phrase was tested in the electronic record (Epic Systems) with several iterations before adopting one to use across centers. During regular meetings, QI CoP members reflected on how they were asking the individuals if they had pain that affects activities and problem solved solutions to do this more consistently. An example provided in one meeting was to ask if they have pain first, which is reflected in the final smart phrase (Figure 2).

**Figure 2:**
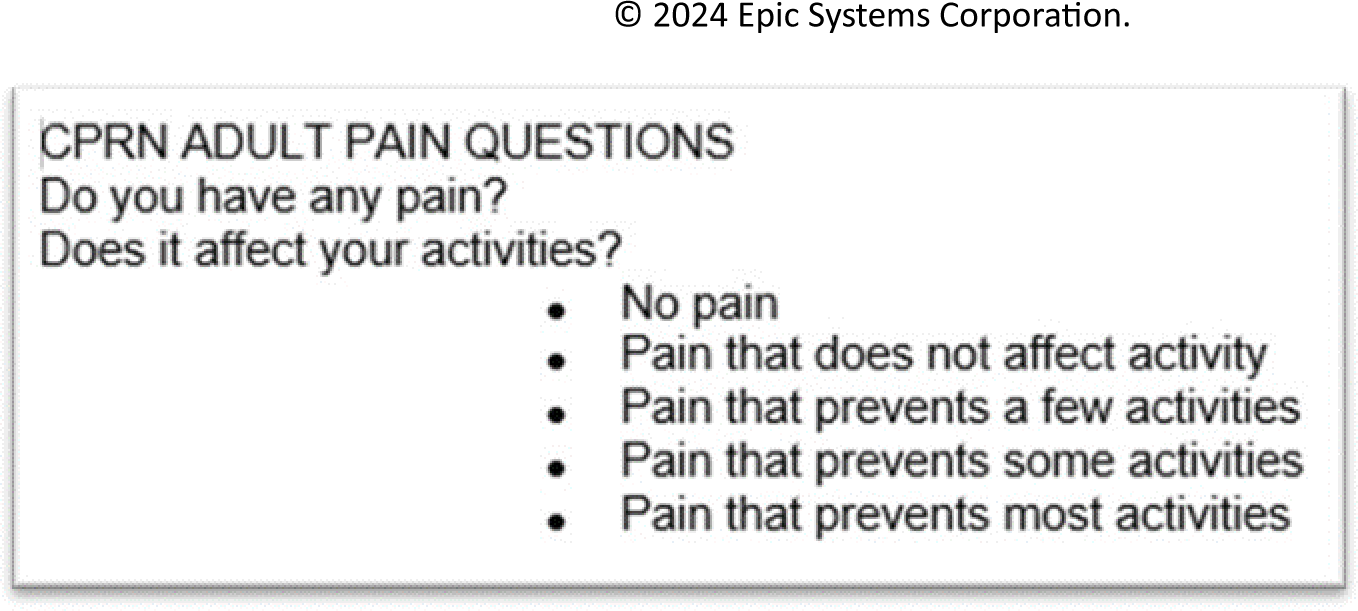
Adopted Smart Phrase for use in electronic medical record documentation. A screen shot showing the section for CPRN centers screening at visits of adults with CP as it appears in the electronic medical record. Do you have any pain? Does it affect your activities? With possible responses as No pain, Pain that does not affect activity, Pain that prevents a few activities, Pain that prevents some activities and Pain that prevents most activities.

#### Give People Access to Information

The QI CoP team had access to information in several ways including via emails and at bimonthly meetings with results of run charts and data checks shared. Further, all documents, recordings of meetings, and presentations were stored on a secured, shared Microsoft Teams page. Giving people access to information allows them to suggest changes, make good decisions, and take action that leads to improvements.[14]

### Analysis

#### Process Measure Analysis

Progress towards achieving the SMART aim (increase the percentage of adult visits screened for pain that affects activities) was measured with a quantitative time series study design. Statistical process control (SPC), which uses techniques to measure and control variation in a process, was applied to monitor the team’s progress toward achieving the SMART aim.[23] Annotated control charts were updated based on data reports from each institution and communicated during biweekly calls. The centerline represented the mean with upper and lower control limits, which are defined in QI methodology at ± 3 standard deviations beyond the mean.[14, 23] Standard criteria (observation of 8 points above or below the centerline) were used to determine whether observed changes in the process measure were due to common cause variation or special cause signifying a change in the measure. Once the process measure goal was met (≥80%) for 6 months, respondents’ answers were collected for analysis. University of Colorado Hospital (UCH) joined the QI CoP in June 2022, and initiated data entry October 2022. Ethics approval was obtained from the University of Hartford for secondary data analysis and the project was deemed exempt from IRB oversight.

#### Characterizing Pain that Affects Activities in Adults with Cerebral Palsy

Responses on the pain attribute of the HUI3 were aggregated among a consecutive sample of visits with four providers from 3/21/2022 - 6/30/2023 at NCH and CUIMC, and from 10/01/2022 - 07/31/2023 at UCH. Descriptive statistics were used to identify measures of central tendency and confidence intervals, grouping respondents by age, GMFCS[12] levels, gender, and responses to the HUI3. Nonparametric analysis of variance and independent sample t-tests evaluated differences between groups. Logistic regression models were used to construct a model to predict responses on the HUI3. Models for the occurrence of pain versus no pain, and pain that affects activity (combining responses for prevents a few, some, or most activities) versus pain that does not affect activities were constructed. Explanatory variables for building the model to predict pain that affects activities included age (continuous), GMFCS level (ordinal), and gender (categorical). Hosmer-Lemeshow Tests evaluated fit for the models with a small Chi square value and large p value indicating a good fit.[24] Sensitivity and specificity greater than 50% indicate the model predicts outcomes accurately better than chance alone. McFadden’s R^2^ values of 0.02 indicate a weak fit and values over 0.40 indicate that a model fits the data very well.[25] SPSS version 29.0 (Chicago, IL) was used for all analysis.

## RESULTS

### Increase Screening for Pain that Affects Activities in Adults with CP

#### Results from Baseline Phase

Prior to initiating improvement interventions, the team manually extracted baseline data by retrospective chart review, sampling all visits of adults with CP of three providers (2 NCH providers and 1 one CUIMC provider) each week between 5/17/21 through 8/20/21. Baseline screening for pain that affects activities was documented in 42% (n =14/33) of visits.

#### Results from Intervention Phase

The intervention phase started on 8/23/21 with the QI CoP meeting regularly twice a month. Initial monitoring started with the three providers (two Developmental Pediatricians at NCH, and a Physiatrist at Columbia) observing the process every 2 weeks with the goal of achieving 80% of adult visits screened for pain that affects activities. Providers at NCH began to use the standardized, EHR-based smart phrase on 9/6/2021 and CUIMC began on 1/10/22.

These efforts resulted in an overall improvement from 42% to 100% over a period of 6 months (3/7/2022). The process continued to be monitored for sustainability every 2 weeks. A small drop in the achievement of the SMART aim was observed but remained over 90% for the remainder of the intervention period (through June 2023). During this period of sustaining our goal, we began collecting/aggregating responses in a convenient sample of visits of adults across centers. In June 2022 an additional provider (Physiatrist from Colorado (UCH)) reviewed the CPRN QI training materials and joined the QI CoP. This providers’ baseline data was 0.

They implemented the smart phrase, began auditing results, and contributing data on 10/3/2022. The final control chart is represented in Figure 3.

**Figure 3:**
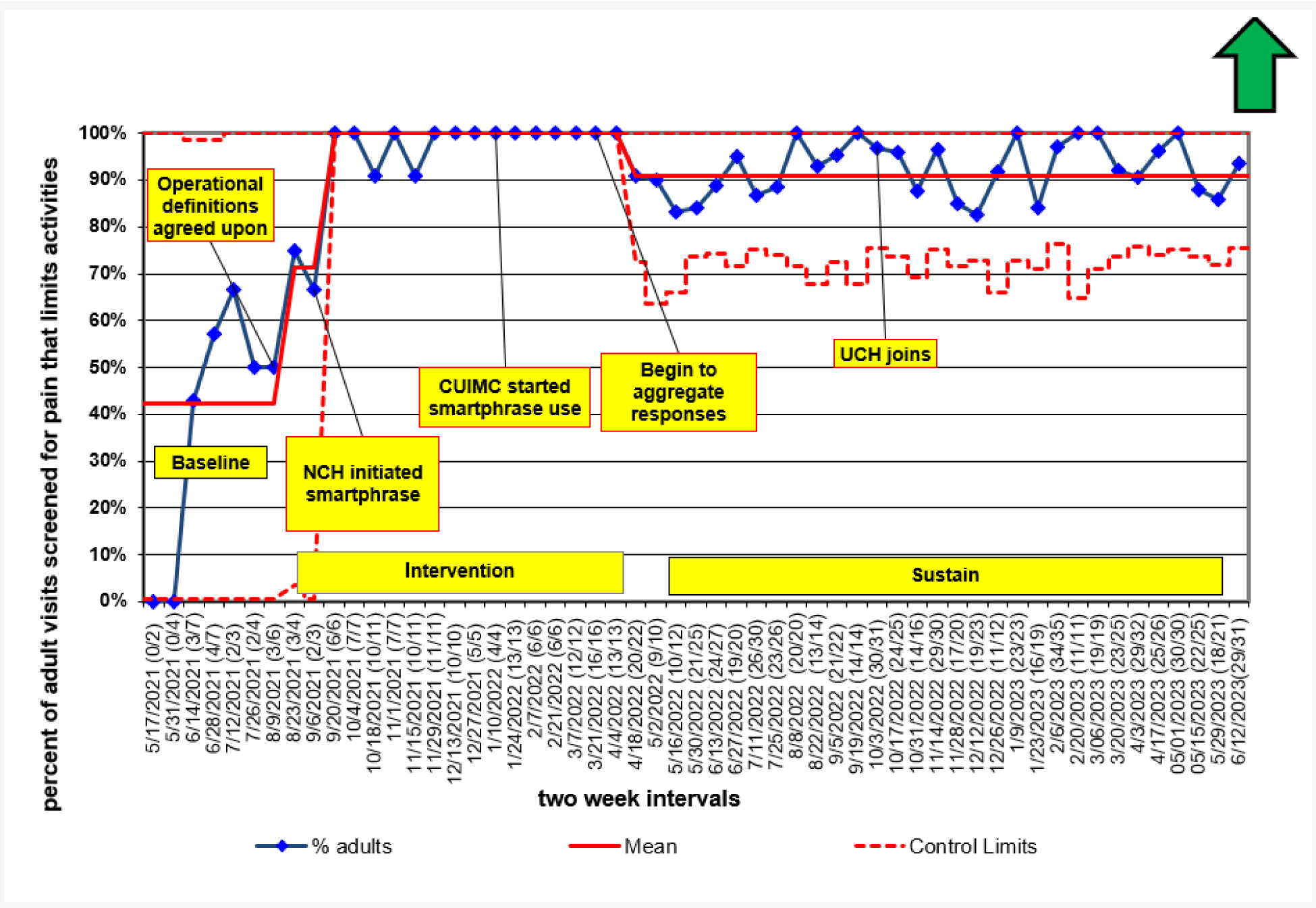
Control Chart Displaying the Progression of the project across time. NCH: Nationwide Children’s Hospital, CUIMC: Columbia University Irving Medical Center, UCH: University of Colorado Hospital. P chart: The percentage of adult CP visits screened for pain that limits activities each 2-week interval. This P chart indicates by 2-week intervals the percentage of adult CP visits over the course of the initiative. The observed values are indicated by the solid lines with markers. The arrow in the upper right-hand corner represents the desired direction of change. The center line is indicated by the solid gray line and represents the overall average score for a specific period. The control limits, indicated by the dashed gray lines are used to determine when the process is out of control. The magnitude of the center line shifts from 42 % to 100% to 91%. A three-section graphical line representation of how many adult visits were screened for pain over three periods (baseline, intervention and sustain). The average number of adult visits screened for pain increased across phases from baseline (42%) to intervention (100%) and dropped slightly but remained over 90% during the sustain phase.

### Characterizing Pain that Affects Activities at Visits of Adults with Cerebral Palsy across 3 Centers

A consecutive sample of visits screened, and the recorded responses were collected from March 2022 - June 2023 for adults receiving care at NCH and CUIMC, and from October 2022 - June 2023 at UCH. Table 1 details the characteristics of 450 adult visits across all 3 centers in the time specified (mean age 31.5 years, range 18-72 years; 49% male, 32% GMFCS V). Ninety-four percent (n=423) of visits were screened for pain that affects activities. One visit was missing responses resulting in 422 visits with data for analysis. Out of 422 visits, 185 (44%, 95% CI 0.39, 0.49) reported pain; and of those with pain, 100/185 (54%, 95% CI 0.49., 0.63) reported it affected activities to some extent.

**Table 1.**
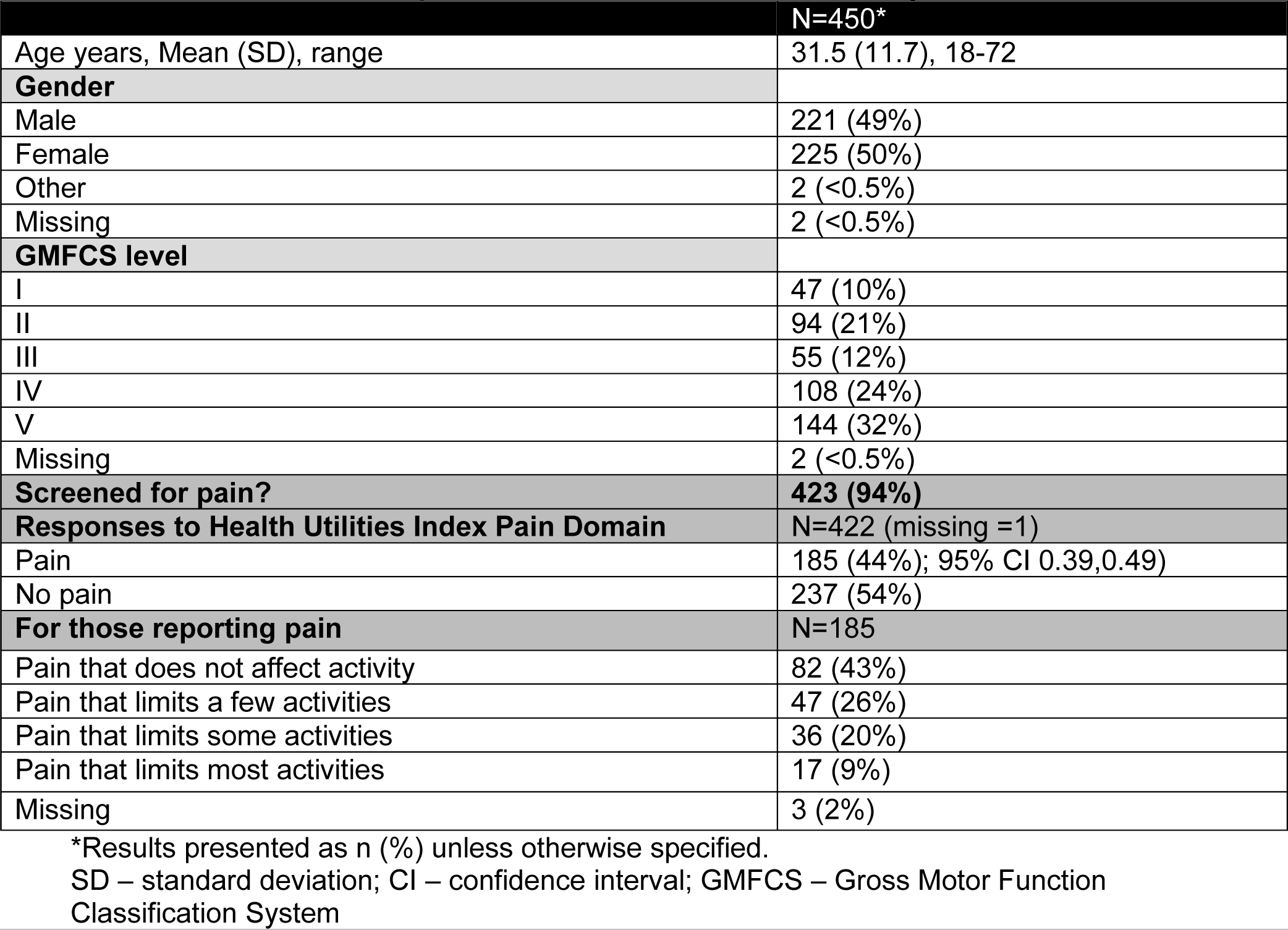
Consecutive Sample of Adult Visits with Cerebral Palsy.

|  | N=450* |
| --- | --- |
| Age years, Mean (SD), range | 31.5 (11.7), 18-72 |
| **Gender** |  |
| Male | 221 (49%) |
| Female | 225 (50%) |
| Other | 2 (<0.5%) |
| Missing | 2 (<0.5%) |
| **GMFCS level** |  |
| I | 47 (10%) |
| II | 94 (21%) |
| III | 55 (12%) |
| IV | 108 (24%) |
| V | 144 (32%) |
| Missing | 2 (<0.5%) |
| **Screened for pain?** | **423 (94%)** |
| **Responses to Health Utilities Index Pain Domain** | N=422 (missing =1) |
| Pain | 185 (44%); 95% CI 0.39,0.49) |
| No pain | 237 (54%) |
| **For those reporting pain** | N=185 |
| Pain that does not affect activity | 82 (43%) |
| Pain that limits a few activities | 47 (26%) |
| Pain that limits some activities | 36 (20%) |
| Pain that limits most activities | 17 (9%) |
| Missing | 3 (2%) |

\*Results presented as n (%) unless otherwise specified.
SD – standard deviation; CI – confidence interval; GMFCS – Gross Motor Function Classification System

### Characteristics of Adults with Pain

Table 2 details the ages, gender, and GMFCS levels of adult visits reporting pain as compared to those without. The visits where pain was reported tended to be older adults (Mann Whitney U = 25871.500, p< .001) and were more often female (X^2^= 35.8, p<0.001) than those without pain. There was no significant difference between the groups in the distribution of GMFCS level, although more than 50% of the adults GMFCS I-III (56% GMFCS I, 54% GMFCS II, and 51% GMFCS III) reported pain as compared to 41% GMFCS IV and 33% GMFCS V.

**Table 2.**
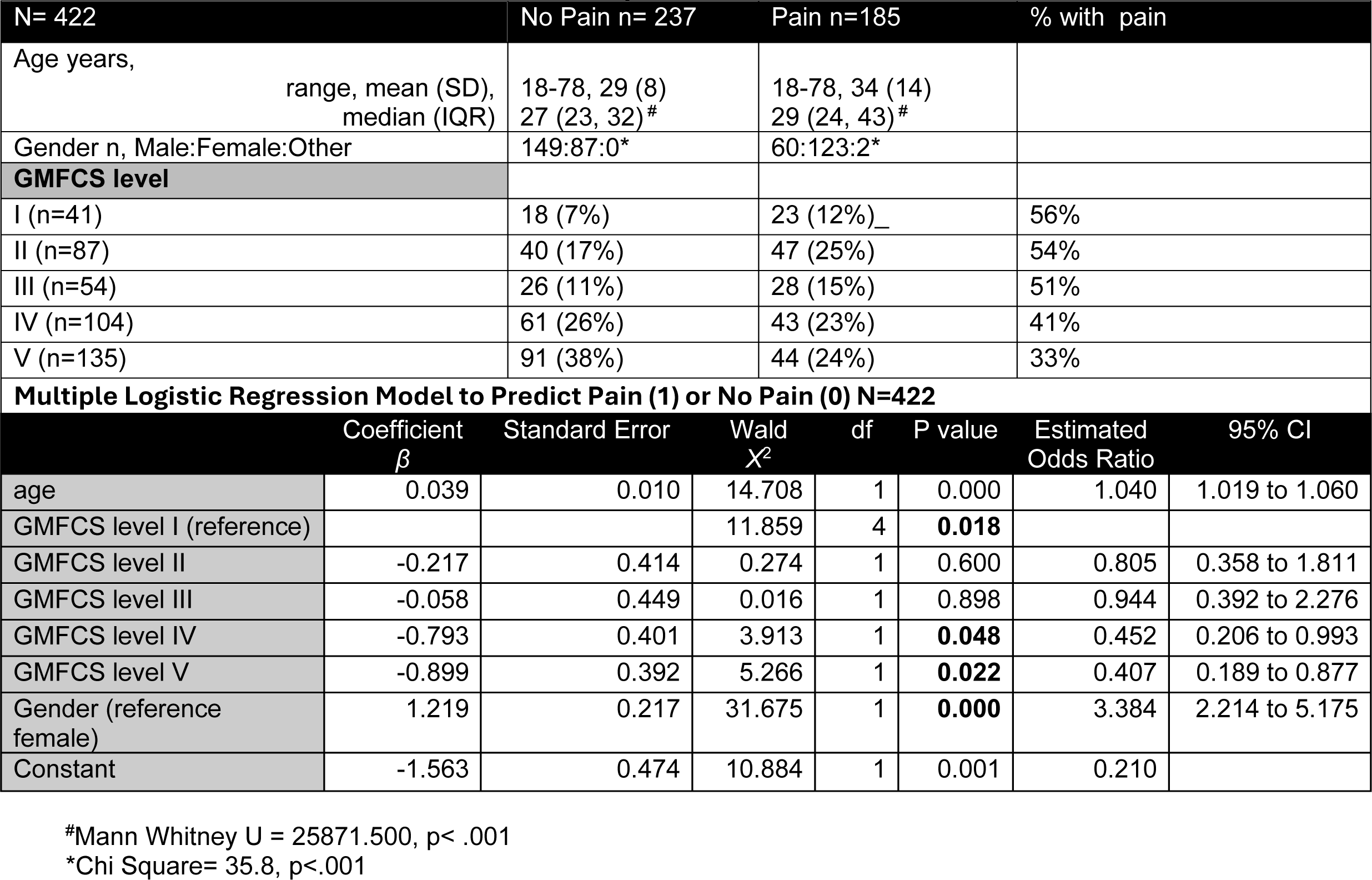
Characteristics of Adult Cerebral Palsy Visits with No Pain and Pain.

A binary logistic regression model was constructed to predict the occurrence of pain based on age, gender, and GMFCS level. (Table 2) The model yielded a R^2^ of 0.20, p<0.001, with Hosmer - Lemenshow Chi Square of 10.0, p=0.264 indicating a good fit.[24] The model predicts who has pain 53% of the time and predicts who does not have pain 81% of the time, with an overall specificity of 69%. Increasing age was associated with a high probability of experiencing pain, with the odds of experiencing pain increasing by 4% for each year (OR=1.04). There was an association of GMFCS level with pain (p=0.018). Individuals classified as GMFCS levels II through V had decreased likelihood of pain relative to those classified as GMFCS I, with GMFCS levels IV and V with the lowest odds of having pain. Gender was associated with pain (p=.000), with females 3.4 times as likely as males to report pain.

### Pain that Affects Activities versus Pain that Does Not Affect Activities

Table 3 describes the characteristics of adult visits reporting pain that affects activities as compared to those with pain that does not affect activities. More females than males reported pain that affects activities, but this did not reach statistical significance (X^2^ = 3.7, p=0.054, 95% CI 0.317, 1.002) nor were there significant differences in age between the groups (t=0.391, p=0.348) or GMFCS levels (Mann Whitney U = 4779, p =0.227). Yet, among adults classified as GMFCS level I, pain that affects activities was reported by 70% as compared to 33% those of GMFCS level IV.

**Table 3.** Characteristics for Visits Reporting Pain that Does not Affect Activities versus Pain that Does Affect Activities.

| N=185<br>Missing:<br>n=3 | Does Not Affect<br>Activities n=82 | Does Affect<br>Activities n=100 | % with pain that affects activity |  |  |  |  |
| --- | --- | --- | --- | --- | --- | --- | --- |
| Age years, range,<br>mean (SD) | 18-72, 34 (12) | 18-71 34, 15 |  |  |  |  |  |
| Gender n,<br>Male:Female:Other | 34:46:2 | 26:74:0 |  |  |  |  |  |
| GMFCS level * |  |  |  |  |  |  |  |
| I (n=23) | 7 (8) | 16 (16) | 70% |  |  |  |  |
| II (n= 50) | 18 (22) | 28 (28) | 56% |  |  |  |  |
| III (n=28) | 12 (15) | 16 (16) | 57% |  |  |  |  |
| IV (n=42) | 28 (34) | 14 (14) | 33% |  |  |  |  |
| V (n=43) | 17 (21) | 26 (26) | 60% |  |  |  |  |
| Multiple Logistic Regression Model to Predict Pain that Affects Activities (1) or Pain that does not Affect Activities at Adult Visits with Pain (N=183) |  |  |  |  |  |  |  |
| | Coefficient<br>$\beta$ | Standard<br>Error | Wald<br>$\chi^2$ | df | P value | Estimated<br>Odds Ratio | 95% CI |
| age | 0.000 | 0.012 | 0.000 | 1 | 0.986 | 1.000 | 0.977 to 1.023 |
| GMFCS level I<br>(reference) |  |  | 11.415 | 4 | **0.022** |  |  |
| GMFCS level II | -0.452 | 0.568 | 0.633 | 1 | 0.426 | 0.637 | 0.209 to 1.937 |
| GMFCS level III | -0.683 | 0.607 | 1.267 | 1 | 0.260 | 0.505 | 0.154 to 1.660 |
| GMFCS level IV | -1.639 | 0.573 | 8.178 | 1 | **0.004** | 0.194 | 0.063 to 0.597 |
| GMFCS level V | -0.450 | 0.562 | 0.641 | 1 | 0.423 | 0.637 | 0.212 to 1.919 |
| Gender<br>(reference<br>female) | 0.803 | 0.339 | 5.611 | 1 | **0.018** | 2.232 | 1.149 to 4.336 |
| Constant | 0.410 | 0.631 | 0.423 | 1 | 0.515 | 1.507 |  |

\*Results presented as n (%) unless otherwise specified.
SD – standard deviation; CI – confidence interval; GMFCS – Gross Motor Function Classification System

A binary logistic regression model was constructed to predict the occurrence of pain that affects activities versus pain that does not affect activities, based on age, gender, and GMFCS. (Table 3) The model yielded a R^2^ of 0.12, p<0.01, with Hosmer Lemenshow Chi Square of 5.85, p=0.664 indicating a good fit. The model predicts who has pain that affects activities 71% of the time and predicts who does not have pain that affects activities 60% of the time, with an overall specificity of 66%. There was an association of GMFCS levels with pain that affects activities (p=0.022). Individuals classified as GMFCS levels II through V had decreased likelihood of pain that affects activities relative to those classified as GMFCS I. Individuals classified as GMFCS levels IV had the lowest odds of having pain that affects activities. Gender was associated with pain (p=.018), with females 2.23 times as likely as males to report pain that affects activities.

## DISCUSSION

This report demonstrates the feasibility of utilizing QI methodology to improve documentation of screening for pain that affects activities during outpatient visits of adults with CP across select providers at three centers within the CPRN. Further, information from real world visits can be utilized to examine factors associated with pain that affects activities. Over approximately one year (8/9/2021-9/19/2022), documentation of screening for pain that affects activities increased from 42% of visits to 91% across three providers from two centers, before spreading to a fourth provider from a third center and maintaining 91% through June 2023. Key drivers of improvement included clinician knowledge about the importance of screening for pain that affects activities, clinician buy-in, sufficient time in the clinic visit, and a standardized process to screen and document that screening occurred. Ongoing meetings and review of the data with the QI CoP allowed for frequent reflection and adaptations to the process. Once the smart aim was achieved, a cohort was identified from all adult CP visits from the four providers’ clinics at three centers, and 44% of visits reported pain. Of these, 66% reported pain that affects activities. In this cohort, older females and those classified as GMFCS level I were more likely to report pain and females GMFCS I were more likely to report pain that affects activities.

The overall mean prevalence of pain in adults with CP has been estimated at 70% with ranges in individual samples between 38% and 89%.[11] Our findings are within that range, but on the lower end. Like the meta-analysis, we identified women at greater risk for reporting pain than men. However, in our sample, adults classified as GMFCS I were more likely to report pain and pain that affects activities. Also in our sample, risk was reduced for pain that affects activities among adults classified at GMFCS IV, while the meta-analysis found risk elevated in both those classified as GMFCS II and IV. These differences are most likely a result of sampling as the meta-analysis[11] only included studies with representative samples and our study sample is biased toward people who receive care at a tertiary care center (whether due to knowledge, access, complexity, or other reasons). For example, our sample included a larger proportion of individuals classified as GMFCS levels IV & V as compared to the sample included in the meta-analysis.

In the meta-analysis[11], the comparison of proxy response to self-report showed minimal differences. In contrast, Rodgy-Bousquet et al[26] reported higher proportion of self-reported pain compared to proxy-reported pain. They also found those of Communication Function Classification System (CFCS)[27] level I reported more pain than those of CFCS levels II-V. We did not distinguish proxy responses from self-report and were unable to do a subgroup analysis. Future work should consider proxy versus self-report responses and examine responses by CFCS levels.

Like the meta-analysis[11], our study also found women were more likely to report pain and that ambulatory adults were more likely to have pain than non-ambulatory adults. Our models for identifying the occurrence of pain and pain that affects activities is weak given the low R^2^ values,[25] but is not surprising as we did not account for other factors that contribute to the pain experience. The International Association of the Study of Pain recommends using a biopsychosocial framework when treating pain, and considering the impacts of anxiety, depression, and social role satisfaction on the pain experience.[28] Access to resources, such as health care, housing, transportation, cultural beliefs, and food security are additional factors that contribute to pain affecting a person’s activity.[28] The current study only used one tool and the meta-analysis used a variety of tools that ask about pain affecting activities in different ways.[11].

A strength of this study was that we were able to routinely incorporate screening during usual clinical care. Conducting this project across centers within CPRN was a strength and demonstrates the impact of QI efforts within a learning health network to change processes and promote patient centered care. Additionally, by involving several centers we were able to identify a larger and more geographically diverse population than at a single center alone. Next steps include automating data extraction, differentiating between proxy and self-report, and distinguishing between chronic versus acute pain. Sustaining these efforts across the three centers will be vital as well as spreading these QI efforts to additional CPRN centers.

### Limitations

QI work is limited by the contextual factors at each institution. The sample is limited by the selection bias of using hospital-based sampling of clinicians who desired to participate and specialized in care of adults with CP. This study only captured what was documented at each of the 94% of visits where documentation was present, and we were not able to know what happened during the 6% of visits where screening for pain that affects activities was not documented. Additionally, our time frame was relatively short and collecting data for 12 months in all three health systems would generate a more accurate period prevalence given variations in visits across the months of the year.[29] Interpretation of the findings is limited by the inclusion of both self- and proxy-report without differentiation, although the HUI3 is used widely with both self- and proxy-report in adult populations.[20] Our sample is biased towards adults with CP of GMFCS levels IV and V. However, the prevalence of pain (44%) is of concern as is the percentage of adults with pain that affects activities.

This study only assessed one important aspect of screening for pain and did not consider other important domains to classify and treat pain such as location and type of pain. QI methodology does not allow us to infer causality of one intervention utilized during the QI process. Rather, QI expects that several interventions will be applied to the process to drive achievement of the smart aim.[14] Different types of pain respond to different types of pain treatments.[30] To effectively optimize treatment plans, the next step is to standardize how pain is classified in adults with CP. Despite these limitations, this is the first study to describe a standardized process to screen for pain that affects activities in adults with CP across multiple centers. This study has provided a unique opportunity to identify a population that would benefit from proactive pain management.

### Next Steps

The QI processes applied in this project will be spread to other CPRN centers. In addition, we will develop consensus on which tools to use to classify pain and use QI methodology to test use of these tools with the goal to improve pain management for adults with CP.

## Data Availability

All data produced in the present study are available upon reasonable request to the authors.

## Acknowledgements

none

